# Multimodal MRI and Machine Learning Uncovers Distinct Progression Patterns in Friedreich Ataxia

**DOI:** 10.64898/2026.04.21.26351375

**Authors:** Susmita Saha, Nellie Georgiou-Karistianis, Vivienne Teo, Louise A. Corben, David J. Szmulewicz, Lachlan T. Strike, Marcondes C. França, Thiago J.R. Rezende, Ian H. Harding

**Affiliations:** School of Psychological Sciences, The Turner Institute for Brain and Mental Health, Monash University, Clayton, Australia; School of Translational Medicine, Monash University, Melbourne, Australia; Bruce Lefroy Centre for Genetic Health Research, Murdoch Children’s Research Institute, Parkville, Victoria, Australia; Department of Paediatrics, University of Melbourne, Parkville, Victoria, Australia; Balance Disorders and Ataxia Service, Royal Victorian Eye and Ear Hospital, East Melbourne, Victoria, Australia; Bionics Department, University of Melbourne, Australia; NeuroMovement Laboratory, Bionics Institute, East Melbourne, Victoria, Australia; Neurology Department, Alfred Hospital, Monash University, Prahran, Victoria, Australia; QIMR Berghofer Medical Research Institute, Brisbane, Queensland, Australia; Department of Neurosciences, School of Medicine, University of São Paulo at Ribeirão Preto (USP-RP), Ribeirão Preto, SP, Brazil

## Abstract

**Background:** Friedreich ataxia (FRDA) is a rare neurodegenerative disorder with substantial heterogeneity in clinical presentation and progression, complicating prognosis and trial design. Neuroimaging offers objective biomarkers to track disease evolution, yet variability in progression patterns remains poorly understood.

**Objective:** To identify biologically meaningful FRDA progression subtypes using longitudinal multimodal MRI and assess their associations with demographic, genetic, and clinical factors.

**Methods:** Longitudinal structural and diffusion MRI data from 54 FRDA and 57 controls were analysed. Annualised progression rates of macrostructural (volumetric) and microstructural (diffusion) features across cerebellum, brainstem, and spinal cord regions were clustered using Gaussian Mixture Models. Cluster robustness was assessed using per-cluster Jaccard similarity and other validation metrics. Random Forest classification examined predictors of cluster membership.

**Results:** Three reproducible clusters/subtypes emerged: micro-dominant/dual progression, characterised by widespread microstructural deterioration with modest volumetric decline; macro-dominant, marked by pronounced volumetric decline with minimal microstructural change; and minimal/no progression, showing negligible change in all measures. FRDA participants predominated in the first two clusters. Random Forest prediction of cluster membership using clinical and demographic variables identified length of the trinucleotide repeat expansion in the *FXN* gene as key predictor.

**Conclusions:** Data□driven clustering of longitudinal MRI identified distinct FRDA subtypes with unique co□progression patterns, underscoring genetic burden as a key driver. Recognising such heterogeneity can improve patient stratification, enable personalised monitoring, and guide targeted therapeutic strategies. Future studies should validate these subtypes in larger, more diverse cohorts and integrate additional biomarkers for enhanced precision.

## INTRODUCTION

Friedreich ataxia (FRDA) is a rare, autosomal recessive neurodegenerative disorder caused primarily by biallelic GAA trinucleotide repeat expansions in the intron 1 of the *FXN* gene (1). Patients experience progressive motor deterioration in the form of ataxia, which is always of cerebellar origin but often contributed to by vestibular and somatosensory impairment. The ataxia manifests widely and includes upper limb functional impairment, loss of ambulation, speech and swallow dysfunction, and ultimately reduced life expectancy. Despite its single-gene origin, FRDA displays clinical heterogeneity with a wide variability in age at symptom onset (AAO), progression rates, clinical phenotypes, and patterns of neurodegeneration (2–6). This variability poses major challenges for prognosis, patient management, therapy and clinical trial design.

Clinical rating scales such as the Friedreich Ataxia Rating Scale (FARS) and the Scale for the Assessment and Rating of Ataxia (SARA) remain the gold-standard tools for monitoring disease progression in FRDA. However, clinical rating scales can be impacted by natural day-to-day variability in symptom severity, patient fatigue, rater subjectivity, and context (7,8). Their sensitivity is greater over longer follow-up periods (≥2 years) than within a single year (9), they are less responsive in late-stage or pediatric cohorts, and their relationship to the underlying biological heterogeneity of progression remains unclear, especially as most studies rely on cross□sectional clinical-imaging associations (10–14).

Neuroimaging offers a complementary strategy for monitoring FRDA by providing objective, quantifiable measures of brain and spinal cord change (13). These measures have shown greater sensitivity than clinical scales for detecting early-stage alterations, particularly over short follow-up periods in other hereditary cerebellar ataxias (15). This capability offers significant potential to accelerate drug development by guiding therapeutic target prioritisation and providing sensitive, quantitative biomarkers for treatment monitoring, participant stratification or recruitment enrichment. The importance of such biomarkers is underscored by regulatory initiatives, including the United States Food and Drug Administration’s (FDA) Biomarker Qualification Program and Critical Path Initiative (16– 18) and the European Medicines Agency’s (EMA) Qualification of Novel Methodologies framework (19).

FRDA neuroimaging studies consistently report macrostructural and microstructural abnormalities using MRI techniques. Volumetric loss is most commonly observed in the cerebellum, brainstem, spinal cord, and cerebellar peduncles, while widespread microstructural changes measured using diffusion-weighted MRI are most notably reported in the superior and inferior cerebellar peduncles, deep cerebellar white matter and peri-dentate regions, dorsal spinal afferent (sensory) pathways, and corticospinal tracts (13,20–24). However, most literature to date has relied on cross□sectional designs, limiting the ability to delineate within□subject trajectories of neurodegeneration. Longitudinal MRI studies are beginning to fill this gap (14,20,21,25,26). For example, a recent study in FRDA reported progressive macrostructural volume loss in cerebellar grey and white matter volumes and microstructural alterations indexed by reduced fractional anisotropy (FA) and increased radial diffusivity (RD) in the superior cerebellar peduncle (SCP), posterior limb of the internal capsule and superior corona radiata over a 36-month period (14). These findings directly support the utility of neuroimaging biomarkers to sensitively quantify longitudinal disease progression in FRDA.

Understanding how genetic and biological factors shape clinical and neuroimaging heterogeneity in FRDA progression remains an active and critical area of research. The size of the smaller expanded allele (GAA1) is inversely correlated with age at onset and time to wheelchair dependence, and both GAA1 and disease duration have been linked to a higher prevalence of complications such as scoliosis, bladder symptoms, impaired vibration and proprioception sense, foot deformity, cardiomyopathy, and diabetes (27–29). Pandolfo (2009) (4) demonstrated that variation in neurophysiological abnormalities correlates with GAA1 repeat size and likely contributes to the heterogeneity of FRDA progression, while neuroinflammation in affected brain regions has been associated with earlier onset but tends to diminish over time (30,31). In addition, Lynch and colleagues (2006) showed that FARS and related performance measures correlated with disability, daily functioning, and disease duration, while age and shorter GAA repeat length predicted FARS scores in site and sex adjusted models (32). Cross□sectional neuroimaging studies further show that volumetric and microstructural abnormalities, particularly within the motor cerebellum (including the dentate nucleus, peri□dentate deep cerebellar white matter, and cerebellar peduncles) as well as the brainstem, demonstrate moderate□to□strong associations with disease severity, duration, and age at onset (22,23). However, inter□individual variability in longitudinal neurodegenerative trajectories remains poorly characterised.

This study addresses this gap by applying a data□driven framework to longitudinal, multimodal MRI alongside clinical, demographic and genetic data. We aim to (i) identify FRDA subtypes based on co-progression patterns across multimodal MRI features of brain and spinal cord, (ii) quantify associations between progression subgroups and individual variables such as clinical measures, demographic factors, genetic markers, and disease history, and (iii) develop an automated model to stratify FRDA individuals into data-driven subgroups based on these variables, supporting future targeted participant selection for clinical trials and improving personalised disease management.

## METHODS

### Dataset Characteristics

This study utilises participant data from studies at Monash University, Melbourne (IMAGE-FRDA, 2013-2016) (21,23), and the University of Campinas, Campinas, Brazil (20). The IMAGE-FRDA study received ethics approval from the Monash Health Human Research Ethics Committee (HREC; project #13201B). The Melbourne cohort included 26 FRDA participants with genetically-confirmed FRDA (homozygous GAA-TR expansion in intron 1 of FXN) and 29 healthy age and sex-matched participants with baseline and two-year follow-up brain and spinal cord T1-weighted MRIs and brain diffusion-weighted MRIs which passed quality-control (QC; see *Feature pre-processing*). The Campinas cohort consisted of 28 genetically confirmed FRDA participants and 28 healthy controls with baseline and one-year follow-up brain and spinal cord structural and diffusion MRI data and adherence to QC criteria. Supplementary Table S1 summarises participant demographics, disease characteristics, genetic profiles, and clinical scores across both datasets. In the Melbourne cohort, full FARS assessments were collected as part of the clinical evaluations, whereas in the Campinas dataset only FARS Part III scores (core neurological items) were available.

### Neuroimaging measures

Macroscopic (volumetric) features of interest included volume of the three cerebellar peduncles, three brainstem segments, and five divisions of the cerebellar cortex; and cross-sectional area and eccentricity of the upper cervical spinal cord. Microstructural (diffusivity) features included fractional anisotropy and three diffusivity measures in the cerebellar peduncles. Full details of all features are provided in Table 1. Detailed MRI acquisition parameters and image processing methods are provided in the Supplementary Methods.

**Table 1.** Neuroimaging, clinical, demographic, and genetic features included in the analysis.

| Domain | Feature(s) | Description |
| --- | --- | --- |
| Structural MRI | SCP, MCP, ICP | Volumes of the superior, middle, and inferior cerebellar peduncles |
|  | Midbrain, Pons, Medulla | Volumes of brainstem regions |
|  | AntCBLM, SupPostCBLM, InfPostCBLM, FlocCBLM, VermisCBLM | Volumes of cerebellar lobes (anterior, superior-posterior, inferior-posterior, flocculonodular, and vermis) |
| Diffusion MRI | FASCP, FAMCP, FAICP | Fractional anisotropy of the superior, middle, and inferior cerebellar peduncles |
|  | MDSCP, MDMCP, MDICP | Mean diffusivity of the superior, middle, and inferior cerebellar peduncles. |
|  | ADSCP, ADMCP, ADICP | Axial diffusivity of the superior, middle, and inferior cerebellar peduncles. |
|  | RDSCP, RDMCP, RDICP | Radial diffusivity of the superior, middle, and inferior cerebellar peduncles. |
| Spinal cord MRI | ECC_C1, ECC_C2 | Eccentricity of the cervical spinal cord at levels C1 and C2. |
|  | CSA_C1, CSA_C2 | Cross-sectional area of the cervical spinal cord at levels C1 and C2. |
| Clinical data | FARS, SARA | Friedreich Ataxia Rating Scale (FARS) and Scale for the Assessment and Rating of Ataxia (SARA). |
| Demographic data | Sex; Age | Sex of participant (male/female); age at baseline visit. |
| Disease History | AAO; Duration | Age at ataxia symptom onset; duration of ataxia symptoms (years) from onset to baseline. |
| Genetic data | GAA1 Repeat length | Length of the expanded GAA trinucleotide repeat in intron 1 of the *FXN* gene. |
| *All imaging measures reflect bilateral measures (e.g., total bilateral volume, or mean bilateral diffusivity)* |  |  |

### Feature pre-processing

Annualised progression rates were computed for each feature in Table 1 according to

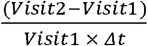

where Δt denotes the inter-scan interval in years.

Participants with >3 extreme outlier features (beyond 3 x interquartile-range) were excluded (n = 2). The influence of study-specific effects due to different scan protocols is mitigated by the within-subject study design (i.e., each subject is their own control). To confirm, site effects were assessed for each feature by testing the significance of a site term in a linear regression model covarying for age, sex (male/female), and group (control/FRDA). No feature demonstrated a significant site effect after multiple-comparison correction (Supplementary Table S2).

### Machine learning clustering

All features were z-scored using standardscaler (mean 0, variance 1) on the combined dataset. Scaled values were used only for model fitting; all descriptive summaries use raw units. We fitted full-covariance Gaussian Mixture Models (GMMs) to the full dataset, including both FRDA and control participants, with the number of clusters (k) ranging from 1 to 15 (random_state = 42, n_init = 5). The optimal cluster number was selected by the Bayesian Information Criterion (BIC). Cluster stability was assessed using 50 bootstrap refits. For each bootstrap iteration, the full dataset (same n) was resampled, a GMM with the optimal k was refit, and cluster labels were predicted. Bootstrap labels were aligned to the base solution using the Hungarian algorithm. Stability was quantified using the Adjusted Rand Index (ARI), Normalised Mutual Information (NMI), and per-cluster Jaccard similarity. Interpretation of these stability metrics and the reported percentile values is provided in Supplementary Note S1. Cluster assignments were subsequently merged back with the raw features to compute per-cluster means and standard deviations. Results were visualised using a feature-by-cluster heatmap and a two-component principal component analysis (PCA) projection to illustrate cluster separation. Cohort composition was additionally summarised by tabulating the distribution of FRDA and control participants across clusters. Within-cluster dispersion was quantified using Mahalanobis distances with a Ledoit-Wolf shrinkage covariance estimator, summarised by cluster and separately for FRDA and control groups (see Supplementary Methods for details). Analyses were performed using Python 3.10.

### Cluster prediction and biological relevance analysis

To explore the biological links of the GMM-derived clusters, we trained a Random Forest classifier to predict cluster membership using age, age at onset (AAO), disease duration, GAA1 repeat length, sex and annual FARS/FARS-III progression rate. Only participants with complete predictor data were included. Sequential forward feature selection was embedded within a nested cross-validation framework, iteratively adding predictors to achieve a maximised balanced accuracy. Hyperparameter optimisation was performed using randomised search within a 3-fold inner cross-validation loop, while stratified 5-fold outer cross-validation generated out-of-fold predictions for unbiased performance estimation. Balanced accuracy was the primary evaluation metric, and per-class recall was additionally reported. The optimal hyperparameter values identified in each outer fold are reported in Supplementary Table S3. Descriptive statistics for all predictors were computed by cluster and visualised using boxplots. Differences across clusters for continuous varaibles were assessed using pairwise Welch’s t-tests. Differences in sex distribution across clusters were assessed using a chi-square test. Benjamini-Hochberg false discovery rate (FDR) correction was applied to account for multiple comparisons.

## RESULTS

### Clusters reveal distinct co-progression patterns across macrostructural and microstructural imaging features

Model selection using the BIC scores identified four clusters as the optimal solution (Figure 1A). The proportion of FRDA participants (relative to controls) in each cluster were as follows: Cluster 1 = 75.0%, Cluster 2 = 31.8%, Cluster 3 = 78.3%, and Cluster 4 = 34.0%. These distributions suggest that Clusters 1 and 3 represent predominantly FRDA-associated progression patterns.

**Figure 1.**
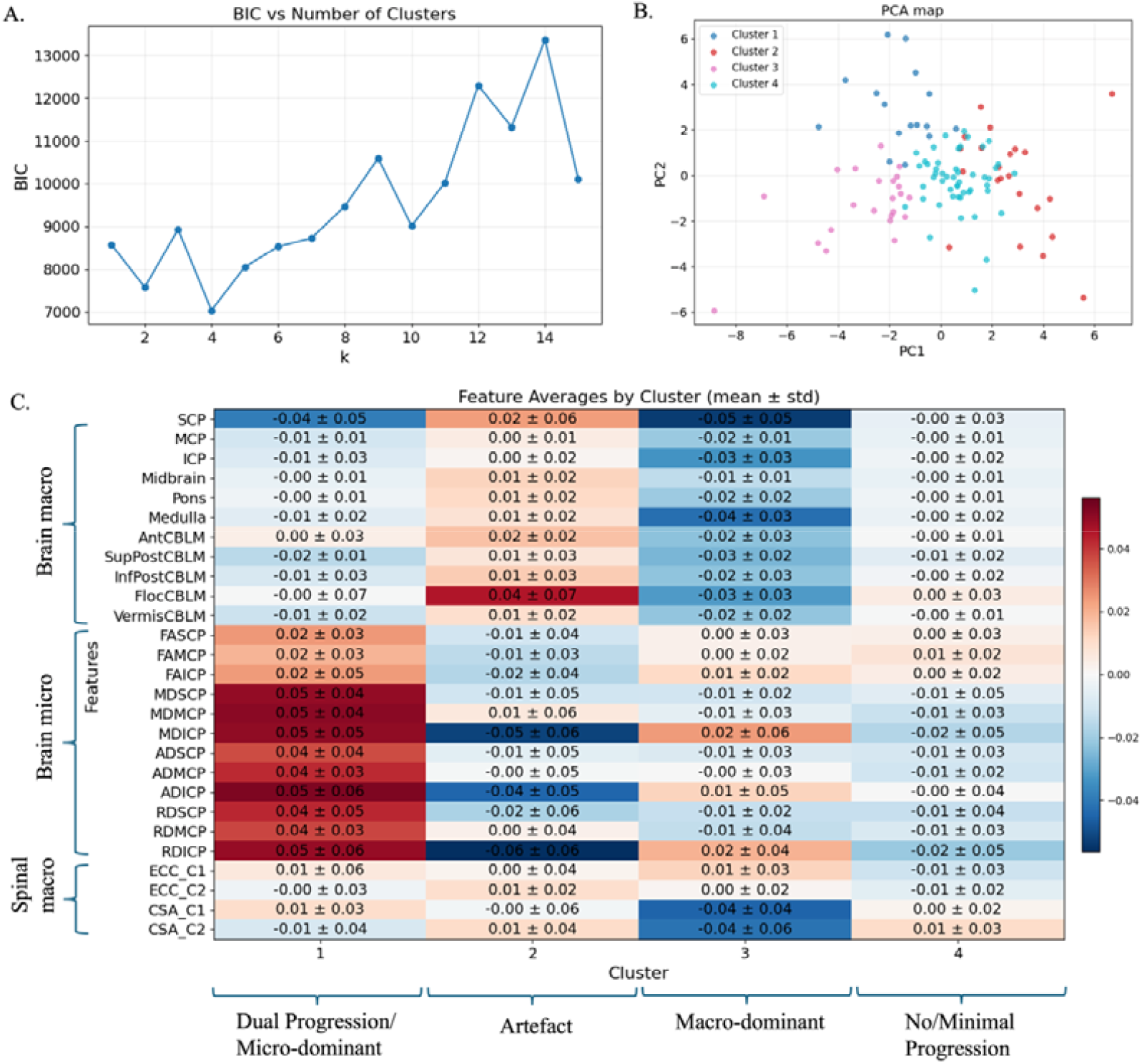
A. BIC scores across tested cluster numbers. B. PCA showing cluster separation. C. Heatmap of annual progression rates for macro- and microstructural features (mean ± SD) in identified clusters; values reflect yearly proportional change (e.g., 0.02 = 2% increase/yr).

Three of the four clusters were biologically interpretable (Figure 1C):

⍰ **Cluster 1 (Dual-progression / micro-dominant):** Characterised by widespread microstructural deterioration accompanied by modest macrostructural atrophy. All of the diffusivity metrics (MD, AD, RD) across all cerebellar peduncles demonstrated positive progression, averaging 2-5%/yr, indicative of microstructural degeneration. Macrostructural change was greatest in SCP volume (4%/yr)), followed by superior-posterior cerebellar lobe volume (2%/yr). Other macrostructural features in the brain and spinal cord showed decreases of 1%/yr or less.
⍰ **Cluster 3 (Macro-dominant):** Defined by volumetric decline across nearly all brain and spinal cord regions (approximately 2-4%/yr in all except midbrain). Microstructural alterations were limited, with increases of 1-2%/yr observed only in inferior cerebellar peduncle diffusion measures.
⍰ **Cluster 4 (Minimal or no progression):** Minimal or no degenerative changes across macro- and microstructural features, consistent with a stable or control-like subtype.

**Cluster 2** was defined by anatomically inconsistent and biologically infeasible effects (i.e., volumetric enlargement, diffusivity decreases), suggesting that this cluster was predominantly capturing noise and measurement variability.

Bootstrap resampling demonstrated moderate-to-good stability and reproducibility of the clustering solution, with stability metrics at the 90th percentile (NMI□□□ = 0.57, ARI□□□ = 0.48, per cluster Jaccard□□□ ≈ 0.67-0.80) for the three biologically interpretable subtypes. Despite modest sample size and uneven cluster distribution (n = 16-50), these values indicate that the core subtype structure was consistently recovered across resamples. Cluster 2 exhibited the weakest stability (Jaccard□□□ = 0.596) (Table 2), in line with the interpretation that it is predominated by noise. Nevertheless, the three dominant clusters showed reliable alignment across bootstrap iterations, supporting the robustness and stability of the derived progression subtypes.

**Table 2.** Cluster composition, summary statistics, and reproducibility indices.

| Cluster | Group | n | % | Shrinkage Mahalanobis Distance |  |  | Jaccard_p90 |
| --- | --- | --- | --- | --- | --- | --- | --- |
|  |  |  |  | Mean | Median | SD |  |
| 1 | CTRL | 4 | 25 | 5.772 | 5.419 | 2.223 | 0.751 |
|  | FRDA | 12 | 75 | 3.905 | 3.828 | 0.499 |  |
| 2 | CTRL | 15 | 68.2 | 5.622 | 5.305 | 2.051 | 0.596 |
|  | FRDA | 7 | 31.8 | 3.18 | 3.294 | 0.546 |  |
| 3 | CTRL | 5 | 21.7 | 5.481 | 4.794 | 1.403 | 0.803 |
|  | FRDA | 18 | 78.3 | 3.712 | 3.696 | 0.805 |  |
| 4 | CTRL | 33 | 66.0 | 4.563 | 4.139 | 1.724 | 0.668 |
|  | FRDA | 17 | 34.0 | 4.13 | 4.338 | 1.075 |  |

PCA visualisation confirmed clear separation among the identified subtypes (Figure 1B). Analysis of within-cluster Mahalanobis distances revealed that FRDA participants formed dense, central cores in clusters 1 and 3, whereas controls were positioned at the periphery (Table 2). The lower mean distances and reduced dispersion among FRDA members indicate that these clusters are predominantly defined by disease traits. However, a small number of controls with partial overlap in some features are also being captured into these classifications, potentially reflecting age-related changes or false positives (supported in part by the greater age of the controls assigned to cluster 3 relative to the no-progression cluster 4; Figure S2). In contrast, cluster 4 displayed minimal differences between FRDA and control participants and exhibited greater overall variability, suggesting a mixed profile which appears to occur not only in healthy individuals but also within a subset of the disease population.

The cluster-wise heatmaps in Figure 2 confirm that the distinct micro-dominant and macro-dominant progression patterns that respectively characterise clusters 1 and 3 on average are also reflected in participant-level profiles. Cluster 4 displays predominantly neutral values across most features for the majority of participants, with only occasional isolated variations, consistent with a minimal or no-progression subtype.

**Figure 2.**
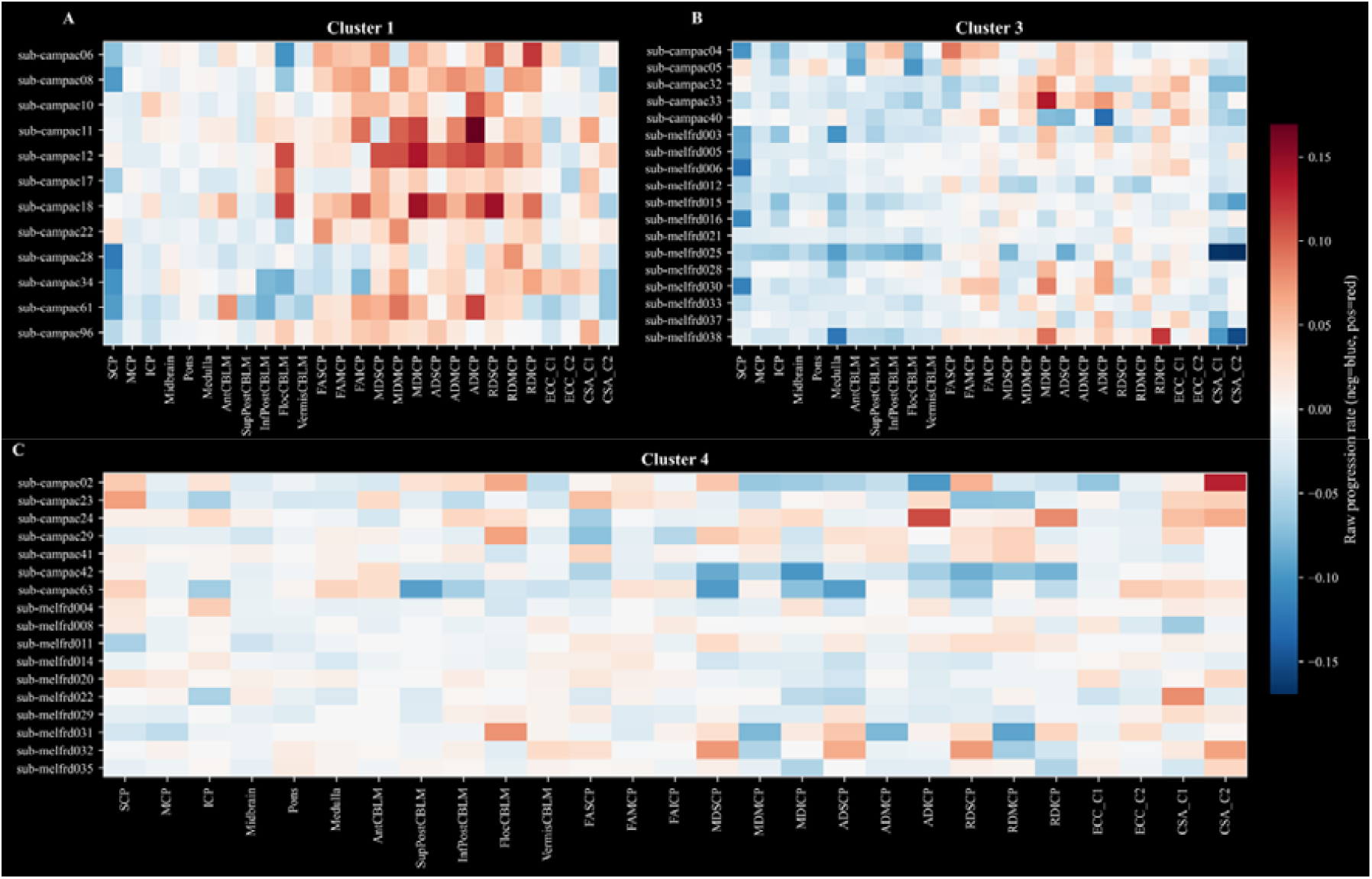
Cluster-wise heatmaps illustrating macro- and micro-feature measures across all FRDA participants, highlighting consistency between cluster-level and individual patterns.

Additional analyses comparing annual progression rates across all imaging features between FRDA participants in each biological cluster (1, 3, and 4) and controls pooled across all clusters further supported distinct cluster-specific progression patterns. In the micro-dominant subtype, only the annual progression rates of diffusion metrics, MD, AD, and RD across all cerebellar peduncles, were significantly different from controls after Welch’s t-tests with Benjamini-Hochberg FDR correction at p < 0.05. No significant differences were observed for fractional anisotropy (FA) or for volumetric measures, except for SCP and MCP. In cluster 2, nearly all volumetric measures of the brain and spinal cord cross-sectional area showed significant progression (p_FDR_<0.05) differences, with the exception of inferior-posterior cerebellar volume. Among the diffusion measures, only RD and MD of the ICP were significantly different. In cluster 3, none of the measures differed significantly between controls and FRDA. These findings further support the cluster interpretation and highlight the distinct progression signatures of the identified subtypes.

### Distribution of demographics, genetic, disease history and clinical variables across clusters

Figure 3 summarises the distributions of genetic (GAA1 repeat length), demographic (age at assessment, sex), clinical (FARS progression rate, annual relative change), and disease-history variables (disease duration, AAO) across the clusters. Among all variables examined, GAA1 repeat length showed the only statistically robust between-cluster differences (Supplementary Table S4). Cluster 1 had significantly greater GAA1 relative to cluster 3 (p_FDR_ = 0.020), and at a trend-level with FDR correction relative to cluster 4 (p_FDR_ = 0.067). GAA1 length was highest in the microstructure-dominant subtype (median = 1055), intermediate in the macrostructure-dominant subtype (median = 685), and lowest in the minimal-progression subtype (median = 614). AAO also showed a trend-level difference between clusters 1 and 4 (p_FDR_ = 0.067). AAO was earliest in the microstructure-dominant subtype (median = 11.5 years) and latest in the minimal-progression subtype (median = 20 years). No significant differences were observed across clusters for age, disease duration, or FARS progression rate.

**Figure 3.**
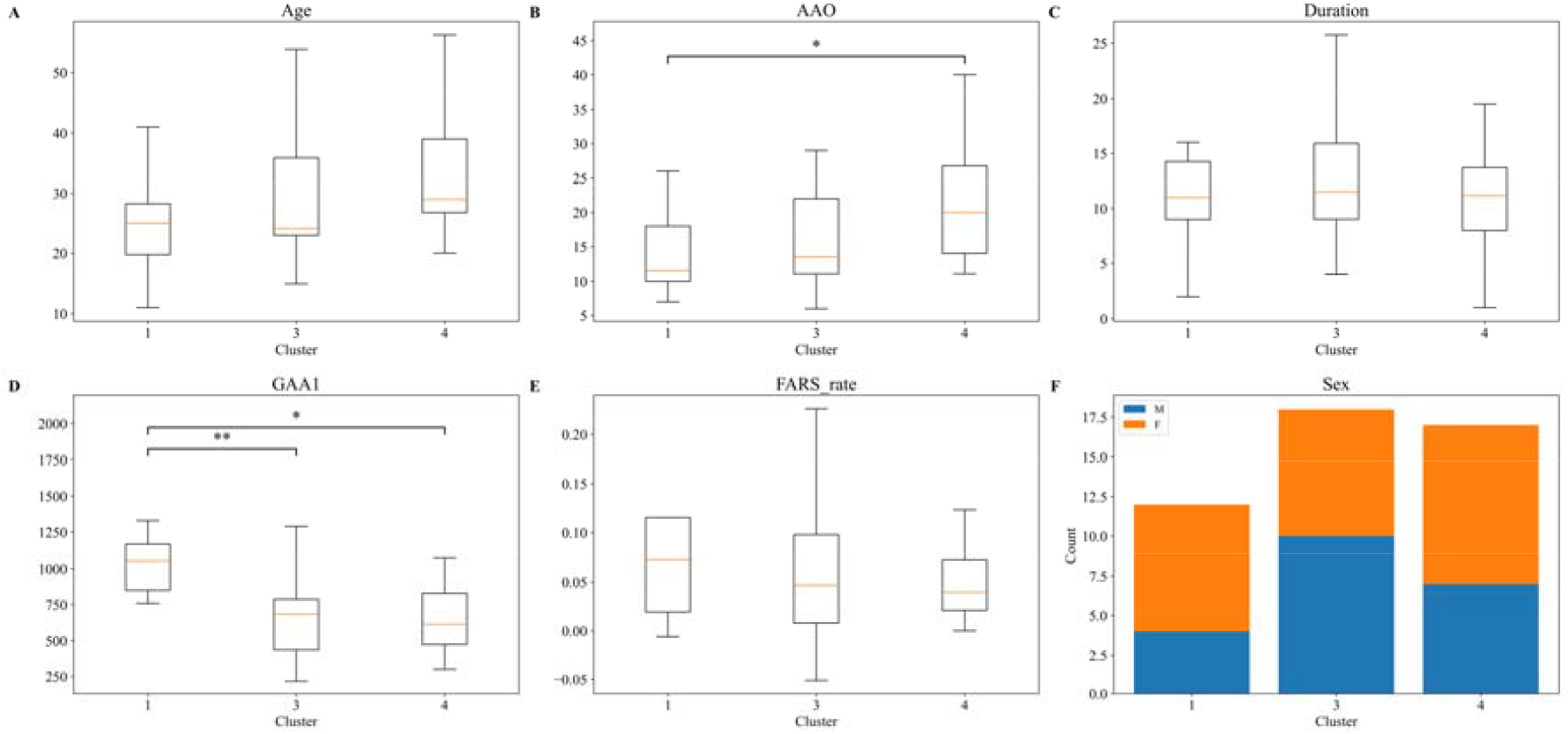
A–E: Boxplots show cluster differences in age (years), AAO (years), disease duration (years), GAA1 repeat length and FARS progression rate (median shown; *p < 0.05 uncorrected; **FDR-corrected). F: Stacked bars show male (M) and female (F) counts per cluster.

### Cluster Prediction from demographics, genetic, disease history and clinical variables

A Random Forest classifier was trained to predict cluster membership of the FRDA participants (Clusters 1, 3, and 4; *n* = 36) using demographic, genetic, disease-history, and clinical variables. Sequential forward feature selection retained only GAA1 repeat length as the optimal predictor. Using nested 5-fold cross-validation, the model achieved a balanced accuracy of 49.2%, exceeding chance level (33%) and indicating modest discriminative performance. Per-class recall varied across clusters: Cluster 1 demonstrated the highest recall (62.5%; *n* = 8), Cluster 3 showed moderate (46.7%; *n* = 15), and Cluster 4 exhibited the lowest performance (38.5%; *n* = 13). These findings suggest that GAA1 repeat length contributes to cluster differentiation but does not fully explain cluster membership. This is consistent with the statistical analyses, in which GAA1 showed the most prominent between-cluster differences, while other variables did not demonstrate robust discriminatory power.

## IV. DISCUSSION

This study characterises heterogeneity in FRDA progression by integrating longitudinal multimodal MRI with data□driven analytical techniques. By leveraging annual macro- and microstructural progression rates across brain and spinal cord regions, we identified three biologically plausible progression subtypes, reflecting distinct co-progression patterns: 1) microstructure-dominant, 2) macrostructure-dominant, and 3) minimal/no progression. These clusters demonstrated technical stability, robust reproducibility, and clear separability in low-dimensional space. Genetic burden (GAA1 repeat length) emerged as the strongest predictor of subtype membership.

In this study, GMMs were chosen for their ability to model complex, multimodal distributions and overlapping phenotypes via probabilistic assignment. Posterior probabilities were largely concentrated in single clusters though, a typical outcome for full-covariance GMMs with modest sample sizes, reflecting high-confidence assignments rather than overfitting, which is appropriate for exploratory subgroup discovery (33). All macro- and microstructural features were retained to avoid bias from pre-selection, as removing correlated features can eliminate contextual interactions critical for disease subtyping (34). Cluster stability, reproducibility, and biological validity were confirmed through complementary validation measures, supporting the methodological approach. In addition, although a small number of controls were also assigned to the FRDA-dominant clusters (1 and 3), controls in cluster 3 tended to be older than those in the minimal-progression cluster (cluster 4) (Supplementary Figure S1), suggesting that age-related neurodegenerative changes may partially explain the overlap with FRDA-associated neuroimaging signatures. Overall, predictive accuracy for cluster membership from demographic, clinical, and genetic variables was modest, with GAA1 repeat length emerged as the only retained predictor. This indicates that genetic burden contributes to cluster differentiation but does not fully explain subtype membership. Classification performance varied across clusters, with the lowest recall observed for the minimal-progression subtype, suggesting greater heterogeneity within this group and the likely contribution of additional biological drivers. Future work should investigate molecular, epigenetic, metabolic, and other factors that may further explain variability in FRDA progression patterns.

Our findings extend previous longitudinal observations from the IMAGE□FRDA cohort, which reported brain region□specific MRI changes in early to mid disease stages (21). We observed 1-3% annual volume reduction across all cerebellar lobules and SCP volume loss in both progression□dominant subtypes, consistent with earlier reports of cerebellar lobule VI and SCP involvement. However, unlike the IMAGE-FRDA study, which reported no significant between-group differences in longitudinal rate of change in diffusion measures (21), our analyses showed that in the microstructure-dominant subtype the progression rates of diffusion metrics (all except FA) were significantly different from controls (p_FDR_<0.05). These findings highlights the value of a data□driven subtyping approach for revealing heterogeneous progression trajectories that may be obscured in traditional group□averaged analyses.

Our predictive modelling and statistical comparisons consistently demonstrated that GAA1 repeat length was the key variable for discriminating the imaging□defined clusters. However, similar cluster□specific trends were observed for both GAA1 and AAO, with the microstructure□dominant subtype showing the highest genetic burden and the earliest onset, while the minimal□progression subtype showing the opposite pattern. This is consistent with the well-established relationship between longer GAA1 and earlier AAO (28,29). A prior longitudinal study by Selvadurai et al. (21) reported a similar association between AAO and microstructural progression in FRDA. Importantly, our clustering approach did not simply capture temporal staging patterns (i.e., participants at different stages of disease progression) described in prior work (22). Instead, the clusters identified in the current work had comparable disease duration, but exhibited heterogeneous patterns of atrophy progression. Cluster□specific differences in GAA1 and AAO are consistent with earlier evidence that these variables influence the rate of clinical progression and risk of ambulation loss (28,29,35). Although FARS progression rates showed only a trend across clusters (cluster 1: 0.07/yr; cluster 3: 0.05/yr; cluster 4: 0.04/yr) and did not reach statistical significance, the pattern is consistent with studies showing that late□onset FRDA is typically associated with smaller repeat expansions and a milder disease course (36,37). Collectively, these findings also align with previous reports demonstrating correlations between neuroimaging progression, clinical progression, and age at disease onset (14,21).

Identifying progression subtypes has key implications for therapy and trial design, and personalised care. Stratifying participants by imaging-derived patterns can reduce cohort heterogeneity, boost statistical power, generate stronger evidence for therapeutic efficacy and enable targeted interventions. These benefits are further amplified where subject recruitment is challenging due to the rarity of the disease of interest, such as is the case with FRDA. Future work should use larger, multi-site cohorts, integrate additional modalities such as functional and susceptibility-based biomarkers, and explore deep learning on raw images for data-driven subtype discovery. Our use of annualised percentage change offers an interpretable index for comparing relative progression across features, facilitating subtype identification. We did not adjust for baseline variability; instead, we captured proportional change relative to each participant’s initial status, reflecting clinically meaningful progression. However, percentage-based measures assume linearity. Future studies should consider absolute or symmetric change metrics, include baseline values, and apply longitudinal trajectory□based models to better capture non□linear and finer□grained progression patterns. Combining these approaches with further investigation of predictive markers for cluster membership will advance precision stratification in clinical trials.

## Data Availability

All data produced in the present study are available upon reasonable request to the authors

## Acknowledgement

The IMAGE-FRDA study was supported by the National Health and Medical Research Council (NHMRC) Project Grant 1184403.

## Data availability statement

Data are available upon reasonable request. Deidentified participant data underlying main results may be provided to researchers who contact the principal investigator of the respective dataset to establish a data-sharing agreement. Approval for data access will be granted on a case-by-case basis at the discretion of the principal investigator.

## Authors’ Roles

Susmita Saha: study conception and design; execution; analysis and interpretation; writing of the manuscript.

Nellie Georgiou-Karistianis: data contributon and interpretation; editing of the manuscript.

Vivienne Teo: execution and data analysis.

David J. Szmulewicz: clinical inputs and editing of the manuscript.

Lachlan T. Strike: data analysis and review of the manuscript.

Marcondes C. França Jr.: data contributon and interpretation

Thiago J.R. Rezende: data contributon and interpretation, review of the manuscript.

Ian H. Harding: analysis and interpretation; project supervision; critical revision and editing of of the manuscript.

## References

1. Campuzano V, Montermini L, Molto MD, Pianese L, Cossée M, Cavalcanti F, et al. Friedreich’s ataxia: autosomal recessive disease caused by an intronic GAA triplet repeat expansion. Science. 1996;271(5254):1423–7.

2. Vavla M, Arrigoni F, Peruzzo D, Montanaro D, Frijia F, Pizzighello S, et al. Functional MRI studies in Friedreich’s Ataxia: a systematic review. Front Neurol. 2022;12:802496.

3. Delatycki MB, Corben LA. Clinical features of Friedreich ataxia. J Child Neurol. 2012 Sep;27(9):1133–7. doi:10.1177/0883073812448230 PubMed PMID: 22752493; PubMed Central PMCID: PMC3674491.

4. Pandolfo M. Friedreich ataxia: the clinical picture. J Neurol. 2009 Mar;256 Suppl 1:3–8. doi:10.1007/s00415-009-1002-3 PubMed PMID: 19283344.

5. Indelicato E, Delatycki MB, Farmer J, França Jr MC, Perlman S, Rai M, et al. A global perspective on research advances and future challenges in Friedreich ataxia. Nat Rev Neurol. 2025;1–12.

6. Rummey C, Corben LA, Delatycki M, Wilmot G, Subramony SH, Corti M, et al. Natural History of Friedreich Ataxia: Heterogeneity of Neurologic Progression and Consequences for Clinical Trial Design. Neurology. 2022 Oct 3;99(14):e1499–510. doi:10.1212/WNL.0000000000200913 PubMed PMID: 35817567; PubMed Central PMCID: PMC9576299.

7. Saute JAM, Donis KC, Serrano-Munuera C, Genis D, Ramirez LT, Mazzetti P, et al. Ataxia rating scales—psychometric profiles, natural history and their application in clinical trials. The Cerebellum. 2012;11(2):488–504.

8. Grobe-Einsler M, Amin AT, Faber J, Völkel H, Synofzik M, Klockgether T. Scale for the assessment and rating of ataxia (SARA): development of a training tool and certification program. The Cerebellum. 2024;23(3):877–80.

9. Friedman LS, Farmer JM, Perlman S, Wilmot G, Gomez CM, Bushara KO, et al. Measuring the rate of progression in Friedreich ataxia: implications for clinical trial design. Mov Disord. 2010;25(4):426–32.

10. Reetz K, Dogan I, Hilgers RD, Giunti P, Parkinson MH, Mariotti C, et al. Progression characteristics of the European Friedreich’s Ataxia Consortium for Translational Studies (EFACTS): a 4-year cohort study. Lancet Neurol. 2021;20(5):362–72.

11. Rummey C, Kichula E, Lynch DR. Clinical trial design for Friedreich ataxia-Where are we now and what do we need? Expert Opin Orphan Drugs. 2018;6(3):219–30.

12. Rummey C, Perlman S, Subramony SH, Farmer J, Lynch DR. Evaluating mFARS in pediatric Friedreich’s ataxia: Insights from the FACHILD study. Ann Clin Transl Neurol. 2024;11(5):1290–300.

13. Georgiou□Karistianis N, Corben LA, Lock EF, Bujalka H, Adanyeguh I, Corti M, et al. Neuroimaging Biomarkers for Friedreich Ataxia: A Cross□Sectional Analysis of the TRACK□FA Study. Ann Neurol. 2025.

14. Adanyeguh IM, Joers JM, Deelchand DK, Hutter DH, Eberly LE, Guo B, et al. Brain MRI detects early-stage alterations and disease progression in Friedreich ataxia. Brain Commun. 2023;5(4):fcad196.

15. Rezende TJ, Petit E, Park YW, Tezenas du Montcel S, Joers JM, DuBois JM, et al. Sensitivity of advanced magnetic resonance imaging to progression over six months in early spinocerebellar ataxia. Mov Disord. 2024;39(10):1856–67.

16. Commissioner O of the. Focus Area: Biomarkers. FDA [Internet]. 2024 Aug 9 [cited 2026 Feb 5]. Available from: https://www.fda.gov/science-research/focus-areas-regulatory-science-report/focus-area-biomarkers

17. Research C for DE and. Medical Imaging and Drug Development. FDA [Internet]. 2020 Jan 31 [cited 2026 Feb 5]. Available from: https://www.fda.gov/drugs/development-resources/medical-imaging-and-drug-development

18. Research C for DE and. FDA [Internet]. FDA; 2025 [cited 2026 Feb 5]. Biomarker Qualification Program. Available from: https://www.fda.gov/drugs/drug-development-tool-ddt-qualification-programs/biomarker-qualification-program

19. Opinions and letters of support on the qualification of novel methodologies for medicine development | European Medicines Agency (EMA) [Internet]. 2011 [cited 2026 Feb 5]. Available from: https://www.ema.europa.eu/en/human-regulatory-overview/research-development/scientific-advice-protocol-assistance/opinions-letters-support-qualification-novel-methodologies-medicine-development

20. Rezende TJ, Silva CB, Yassuda CL, Campos BM, D’Abreu A, Cendes F, et al. Longitudinal magnetic resonance imaging study shows progressive pyramidal and callosal damage in Friedreich’s ataxia. Mov Disord. 2016;31(1):70–8.

21. Selvadurai LP, Georgiou-Karistianis N, Shishegar R, Sheridan C, Egan GF, Delatycki MB, et al. Longitudinal structural brain changes in Friedreich ataxia depend on disease severity: the IMAGE-FRDA study. J Neurol. 2021;268:4178–89.

22. Harding IH, Chopra S, Arrigoni F, Boesch S, Brunetti A, Cocozza S, et al. Brain structure and degeneration staging in Friedreich ataxia: magnetic resonance imaging volumetrics from the ENIGMA□ataxia working group. Ann Neurol. 2021;90(4):570–83.

23. Selvadurai LP, Corben LA, Delatycki MB, Storey E, Egan GF, Georgiou□Karistianis N, et al. Multiple mechanisms underpin cerebral and cerebellar white matter deficits in Friedreich ataxia: The IMAGE□FRDA study. Hum Brain Mapp. 2020;41(7):1920–33.

24. Rezende TJ, Adanyeguh IM, Arrigoni F, Bender B, Cendes F, Corben LA, et al. Progressive spinal cord degeneration in Friedreich’s ataxia: results from ENIGMA□ataxia. Mov Disord. 2023;38(1):45–56.

25. Joers JM, Adanyeguh IM, Deelchand DK, Hutter DH, Eberly LE, Iltis I, et al. Spinal cord magnetic resonance imaging and spectroscopy detect early-stage alterations and disease progression in Friedreich ataxia. Brain Commun. 2022;4(5):fcac246.

26. Mascalchi M, Toschi N, Giannelli M, Ginestroni A, Della Nave R, Tessa C, et al. Regional cerebral disease progression in Friedreich’s ataxia: a longitudinal diffusion tensor imaging study. J Neuroimaging. 2016;26(2):197–200.

27. Parkinson MH, Boesch S, Nachbauer W, Mariotti C, Giunti P. Clinical features of Friedreich’s ataxia: classical and atypical phenotypes. J Neurochem. 2013;126:103–17.

28. Dürr A, Cossee M, Agid Y, Campuzano V, Mignard C, Penet C, et al. Clinical and genetic abnormalities in patients with Friedreich’s ataxia. N Engl J Med. 1996;335(16):1169–75.

29. Filla A, De Michele G, Cavalcanti F, Pianese L, Monticelli A, Campanella G, et al. The relationship between trinucleotide (GAA) repeat length and clinical features in Friedreich ataxia. Am J Hum Genet. 1996;59(3):554.

30. Apolloni S, Milani M, D’Ambrosi N. Neuroinflammation in Friedreich’s ataxia. Int J Mol Sci. 2022;23(11):6297.

31. Khan W, Corben LA, Bilal H, Vivash L, Delatycki MB, Egan GF, et al. Neuroinflammation in the cerebellum and brainstem in Friedreich ataxia: an [18F]□FEMPA PET study. Mov Disord. 2022;37(1):218–24.

32. Lynch D, Farmer J, Tsou A, Perlman S, Subramony S, Gomez C, et al. Measuring Friedreich ataxia: complementary features of examination and performance measures. Neurology. 2006;66(11):1711–6.

33. Selvadurai LP, Georgiou-Karistianis N, Shishegar R, Sheridan C, Egan GF, Delatycki MB, et al. Longitudinal structural brain changes in Friedreich ataxia depend on disease severity: the IMAGE-FRDA study. J Neurol. 2021;268(11):4178–89.

34. Selvadurai LP, Harding IH, Corben LA, Stagnitti MR, Storey E, Egan GF, et al. Cerebral and cerebellar grey matter atrophy in Friedreich ataxia: the IMAGE-FRDA study. J Neurol. 2016;263(11):2215–23.

35. McLachlan G. Finite mixture models. Wiley-Intersci Publ. 2000.

36. Sirocchi C, Urschler M, Pfeifer B. Feature graphs for interpretable unsupervised tree ensembles: centrality, interaction, and application in disease subtyping. BioData Min. 2025;18(1):15.

37. Mateo I, Llorca J, Volpini V, Corral J, Berciano J, Combarros O. Expanded GAA repeats and clinical variation in Friedreich’s ataxia. Acta Neurol Scand. 2004;109(1):75–8.

38. Lecocq C, Charles P, Azulay J, Meissner W, Rai M, N’Guyen K, et al. Delayed□onset Friedreich’s ataxia revisited. Mov Disord. 2016;31(1):62–9.

39. Patel M, Isaacs CJ, Seyer L, Brigatti K, Gelbard S, Strawser C, et al. Progression of Friedreich ataxia: quantitative characterization over 5 years. Ann Clin Transl Neurol. 2016;3(9):684–94.

