## Supplementary File for "Multimodal MRI and Machine Learning Uncovers Distinct Progression Patterns in Friedreich Ataxia"

**SUPPLEMENTARY METHODS**

**Neuroimaging measures**

For the IMAGE-FRDA study (Melbourne) (1,2), whole-brain T1-weighted magnetization-prepared rapid gradient echo (MPRAGE) and diffusion-weighted MRI scans were obtained using 3-Tesla Siemens Skyra (Siemens, Erlangen, Germany) with a 32-channel head coil. T1-weighted images were acquired with Turbo flash sequence over 4 min and 26s with 176 sagittal slices, voxel size 1 × 1 × 1 mm^3^, matrix size  256 × 256, field of view 256 × 256 mm^2^ and echo time/repetition time (TE/TR) of 2.55ms/1540 ms. Diffusion-weighted images were acquired with spin-echo echo-planar imaging (SE-EPI) sequence over 2 min and 28s using an echo-planar spin-echo sequence, with b = 1200 s/mm², 12 gradient directions, 50 axial slices, voxel size  2 × 2 × 2 mm3, matrix size 128 × 128, field of view 256 × 256 mm^2^ and TE/TR of 100ms/9.8s.

In the Campinas study (3), whole-brain T1-weighted and diffusion-weighted MRIs were acquired using a 3-Tesla Achieva-Intera Philips scanner (Philips, Best, The Netherlands) equipped with an 8‑channel head coil. High-resolution spoiled gradient-recalled echo T1-weighted images were obtained in sagittal orientation with voxel size 1 × 1 × 1 mm³, matrix size 240 × 240 × 180, TE/TR 3.201ms/7 ms, and flip angle 8°. Diffusion-weighted images were acquired over approximately 6 min using a spin-echo DTI sequence, with b = 1000 s/mm², 32 gradient directions, no signal averaging, 70 slices, voxel size 2 × 2 × 2 mm³ (interpolated to 1 × 1 × 2 mm³), matrix size 256 × 256, and TE/TR 61/8,500 ms.

*Structural MRI*

Cerebellar peduncle volumes were estimated using the *segment* function of the Computational Anatomy Toolbox (CAT) v12.9 (4) for SPM12 (<http://www.fil.ion.ucl.ac.uk/spm>), with regions defined using a probabilistic white matter atlas (5) as previously described. Brainstem and cerebellar/cerebral structure volumes were estimated using the automated tools FreeSurfer v7.4.1 (6,7) and FastSurfer v2.3.3 (including the CerebNet cerebellum segmentation algorithm (8,9), respectively.

*Diffusion MRI*

Diffusion-weighted MRI data were preprocessed using QSIprep v1.0.0rc2 (10). Whole-brain diffusion metrics (i.e., FA, AD, MD, RD) were calculated using DIPY v1.9.0 (11). Diffusion metrics were extracted for regions of interest (ROIs) following the ENIGMA-DTI protocol (12), with ROIs defined by the JHU white matter atlas (13).

*Spinal Cord MRI*

Eccentricity and cross-sectional area of the upper cervical spinal cord at vertebral levels C1 and C2, were derived using the ENIGMA-SC pipeline (14) based on the Spinal Cord Toolbox (15) and quality-controlled with the QC_SpinalCord tool (16).

**Within-cluster dispersion analysis**

To characterise within-cluster dispersion, Mahalanobis distances ($d)$ were computed using a Ledoit–Wolf shrinkage covariance estimator (17):

$d_{i}= \sqrt{{(x-\mu)}^{T}\sum_{LW}^{-1} (x_{i}-\mu)}$,

where $x_{i}$ denotes the feature vector for participant i, μ the cluster-specific mean vector and Σ_LW_ the Ledoit–Wolf covariance matrix, estimated separately for each cluster using all assigned participants. Because sample covariance estimates can become unstable when features are correlated or sample sizes are modest, we used the Ledoit–Wolf estimator to obtain a regularised, positive‑definite covariance matrix with a stable inverse, enabling reliable computation of Mahalanobis distances.

**SUPPLEMENTARY FIGURES**

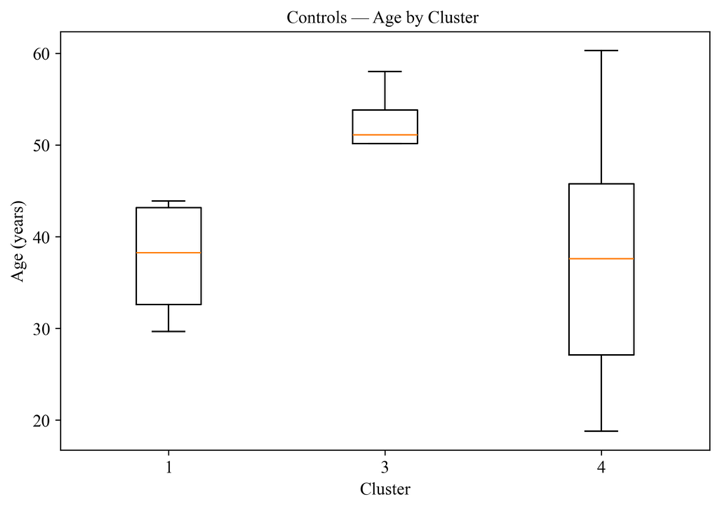

**Figure S1: Age distribution of control participants across clusters.**

The central line indicates the median, boxes represent the interquartile range (IQR), and whiskers extend to 1.5 × IQR.

**SUPPLEMENTARY TABLES**

Table S1: Demographics of Melbourne and Campinas participants included in the study.

| Site | FRDA | | | | | | | Control | |
| --- | --- | --- | --- | --- | --- | --- | --- | --- | --- |
|  | Sex (F/M) | Age (years) | AAO (years) | Duration (years) | GAA1 | FARS_  v1 | FARS_  v2 | Sex (F/M) | Age  (years) |
| Melbourne | 11/15 | 35.46 ± 12.15 | 19.73 ± 9.44 | 15.61 ± 7.30 | 556.92 ± 236.30 | 79.19 ± 26.37 | 86.01 ± 26.71 | 14/15 | 38.55 ± 13.76 |
| Campinas | 17/11 | 26.82 ± 9.53 | 16.19 ± 7.56^a^ | 10.82 ± 6.57 | 997.61 ± 285.88^b^ | 55.80 ± 18.01 | 64.61 ± 17.64^c^ | 20/8 | 39.98 ± 8.50 |
| Total | 28/26 | 30.98 ± 11.61 | 17.92 ± 8.64 | 13.13 ± 7.28 | 737.20 ± 335.88 | 67.28 ± 25.22 | 76.97 ± 25.43 | 34/23 | 39.25 ± 11.40 |

* *Values are presented as mean ± standard deviation. AAO = age at ataxia onset.*

ᵃ Age at onset (AAO) available for 53 participants (Melbourne n=26; Campinas n=27). ᵇ GAA1 repeat length available for 44 participants (Melbourne n=26; Campinas n=18). ᶜ FARS v2 available for 45 participants (Melbourne n=26; Campinas n=19).

Table S2: Statistical assessment of site effects across imaging features

|  | **t value** | **p value** | **p value fdr** |
| --- | --- | --- | --- |
| ADICP | 0.74 | 0.46 | 0.79 |
| ADMCP | -1.09 | 0.28 | 0.66 |
| ADSCP | -0.82 | 0.41 | 0.75 |
| AntCBLM | -1.43 | 0.16 | 0.66 |
| CSA_C1 | -0.67 | 0.50 | 0.81 |
| CSA_C2 | -0.29 | 0.78 | 0.93 |
| CSA_C3 | -0.28 | 0.78 | 0.93 |
| ECC_C1 | 0.00 | 1.00 | 1.00 |
| ECC_C2 | -0.97 | 0.34 | 0.68 |
| ECC_C3 | 1.05 | 0.30 | 0.66 |
| FAICP | 0.14 | 0.89 | 0.99 |
| FAMCP | -0.42 | 0.68 | 0.89 |
| FASCP | -0.43 | 0.66 | 0.89 |
| FlocCBLM | -2.39 | 0.02 | 0.42 |
| ICP | -1.30 | 0.20 | 0.66 |
| InfPostCBLM | -1.14 | 0.26 | 0.66 |
| MCP | -0.06 | 0.95 | 1.00 |
| MDICP | 0.25 | 0.80 | 0.93 |
| MDMCP | -1.41 | 0.16 | 0.66 |
| MDSCP | -0.54 | 0.59 | 0.86 |
| Medulla | -2.22 | 0.03 | 0.42 |
| Midbrain | -1.16 | 0.25 | 0.66 |
| Pons | -2.00 | 0.05 | 0.42 |
| RDICP | 1.20 | 0.23 | 0.66 |
| RDMCP | -0.93 | 0.35 | 0.68 |
| RDSCP | 0.05 | 0.96 | 1.00 |
| SCP | -1.92 | 0.06 | 0.42 |
| SupPostCBLM | -1.29 | 0.20 | 0.66 |
| VermisCBLM | 0.55 | 0.58 | 0.86 |

Values shown correspond to the t-statistic for the site term, the uncorrected p-value, and the false discovery rate (FDR)-adjusted p-value (Benjamini-Hochberg correction)

Table S3: Random Forest Hyperparameters

| Fold | n_estimators | min_samples_split | min_samples_leaf | max_features | max_depth |
| --- | --- | --- | --- | --- | --- |
| 1 | 180 | 6 | 3 | None | 12 |
| 2 | 120 | 6 | 1 | √p | 12 |
| 3 | 180 | 6 | 3 | None | 12 |
| 4 | 240 | 4 | 1 | None | None |
| 5 | 240 | 6 | 1 | √p | 6 |

n_estimators: Number of trees in the forest; min_samples_split: Minimum number of samples required to split an internal node; min_samples_leaf: Minimum number of samples required at a leaf node; max_features: Number of predictors considered at each split (√p indicates the square root of the total number of predictors; *None* indicates that all available predictors were considered); max_depth: Maximum depth of each decision tree (*None* indicates that trees were grown without an explicit depth limit).

### Table S4. Summary of variables showing significant differences between clusters (p < 0.05).

| Feature | Cluster Comparison | p-value (unadjusted) | p-value (FDR-adjusted) |
| --- | --- | --- | --- |
| AAO | 1 vs 4 | 0.011* | 0.067 |
| GAA1 | 1 vs 3 | 0.001* | 0.020* |
| GAA1 | 1 vs 4 | 0.013* | 0.067 |

Significance is indicated by an asterisk (*).

**SUPPLEMENTARY NOTE S1. Interpretation of clustering stability metrics**

Cluster stability was evaluated using bootstrap resampling and quantified using three complementary similarity metrics: the Adjusted Rand Index (ARI), Normalised Mutual Information (NMI), and per-cluster Jaccard similarity. These metrics quantify the agreement between clustering assignments obtained from bootstrap resampling and the reference clustering solution.

The Adjusted Rand Index (ARI) measures how consistently pair of observations are assigned to the same or different clusters in the two clustering solutions, adjusting for chance agreement. Values range from −1 to 1, where 1 indicates identical clustering, 0 corresponds to random agreement, and negative values indicate less agreement than expected by chance (18).

Normalised Mutual Information (NMI) measures the amount of shared information between two clustering solutions, quantifying how much knowing the cluster assignment in one solution reduces uncertainty about the assignment in the other. Unlike ARI, which is based on pairwise agreement of observations, NMI evaluates how well the overall clustering structure is preserved. Values range from 0 (no shared information) to 1 (perfect agreement) (19).

The Jaccard similarity coefficient measures the overlap in membership of individual clusters between two clustering solutions. It is defined as the ratio of the intersection to the union of cluster memberships and ranges from 0 (no overlap) to 1 (perfect overlap) (20).

The values reported in the main text correspond to the 90th percentile (p90) of the bootstrap similarity distribution. This percentile indicates the similarity value below which 90% of bootstrap results fall, representing the level of agreement achieved in the most stable resamples. Reporting the 90th percentile provides an estimate of clustering reproducibility under favourable resampling conditions while remaining robust to occasional low-similarity bootstrap solutions, especially in datasets with small sample sizes.

**REFERENCES**

1.	Selvadurai LP, Georgiou-Karistianis N, Shishegar R, Sheridan C, Egan GF, Delatycki MB, et al. Longitudinal structural brain changes in Friedreich ataxia depend on disease severity: the IMAGE-FRDA study. J Neurol. 2021;268:4178–89.

2.	Selvadurai LP, Corben LA, Delatycki MB, Storey E, Egan GF, Georgiou‐Karistianis N, et al. Multiple mechanisms underpin cerebral and cerebellar white matter deficits in Friedreich ataxia: The IMAGE‐FRDA study. Hum Brain Mapp. 2020;41(7):1920–33.

3.	Rezende TJ, Silva CB, Yassuda CL, Campos BM, D’Abreu A, Cendes F, et al. Longitudinal magnetic resonance imaging study shows progressive pyramidal and callosal damage in Friedreich’s ataxia. Mov Disord. 2016;31(1):70–8.

4.	Gaser C, Dahnke R, Thompson PM, Kurth F, Luders E, Alzheimer’s Disease Neuroimaging Initiative. CAT: a computational anatomy toolbox for the analysis of structural MRI data. Gigascience. 2024;13:giae049.

5.	Van Baarsen K, Kleinnijenhuis M, Jbabdi S, Sotiropoulos SN, Grotenhuis J, van Walsum A van C. A probabilistic atlas of the cerebellar white matter. Neuroimage. 2016;124:724–32.

6.	Iglesias JE, Augustinack JC, Nguyen K, Player CM, Player A, Wright M, et al. A computational atlas of the hippocampal formation using ex vivo, ultra-high resolution MRI: Application to adaptive segmentation of in vivo MRI. Neuroimage. 2015;115:117–37.

7.	Fischl B. FreeSurfer. Neuroimage. 2012;62(2):774–81.

8.	Faber J, Kügler D, Bahrami E, Heinz LS, Timmann D, Ernst TM, et al. CerebNet: A fast and reliable deep-learning pipeline for detailed cerebellum sub-segmentation. NeuroImage. 2022 Dec 1;264:119703. doi:10.1016/j.neuroimage.2022.119703

9.	Henschel L, Conjeti S, Estrada S, Diers K, Fischl B, Reuter M. FastSurfer - A fast and accurate deep learning based neuroimaging pipeline. NeuroImage. 2020 Oct 1;219:117012. doi:10.1016/j.neuroimage.2020.117012

10.	Cieslak M, Cook PA, He X, Yeh FC, Dhollander T, Adebimpe A, et al. QSIPrep: an integrative platform for preprocessing and reconstructing diffusion MRI data. Nat Methods. 2021 Jul 1;18(7):775–8. doi:10.1038/s41592-021-01185-5

11.	Garyfallidis E, Brett M, Amirbekian B, Rokem A, Van Der Walt S, Descoteaux M, et al. Dipy, a library for the analysis of diffusion MRI data. Front Neuroinformatics. 2014;8:8.

12.	Kochunov P, Hong LE, Dennis EL, Morey RA, Tate DF, Wilde EA, et al. ENIGMA‐DTI: Translating reproducible white matter deficits into personalized vulnerability metrics in cross‐diagnostic psychiatric research. Hum Brain Mapp. 2022;43(1):194–206.

13.	Mori S, Oishi K, Jiang H, Jiang L, Li X, Akhter K, et al. Stereotaxic white matter atlas based on diffusion tensor imaging in an ICBM template. NeuroImage. 2008 Apr 1;40(2):570–82. doi:10.1016/j.neuroimage.2007.12.035

14.	Rezende TJ, Adanyeguh IM, Arrigoni F, Bender B, Cendes F, Corben LA, et al. Progressive spinal cord degeneration in Friedreich’s ataxia: results from ENIGMA‐ataxia. Mov Disord. 2023;38(1):45–56.

15.	De Leener B, Lévy S, Dupont SM, Fonov VS, Stikov N, Collins DL, et al. SCT: Spinal Cord Toolbox, an open-source software for processing spinal cord MRI data. Neuroimage. 2017;145:24–43.

16.	[Rensonnet G, Crona N. QC_SpinalCord: Automated quality control for spinal cord MRI [Internet]. [cited 2025 Nov 24]. Available from: https://github.com/art2mri/QC_SpinalCord?tab=readme-ov-file](https://www.zotero.org/google-docs/?pz46JH)

17.	Ledoit O, Wolf M. A well-conditioned estimator for large-dimensional covariance matrices. J Multivar Anal. 2004;88(2):365–411.
